# Estimating vaccine confidence levels among future healthcare workers and their trainers: A quantitative study protocol

**DOI:** 10.1101/2021.02.03.21251068

**Authors:** Elizabeth O. Oduwole, Hassan Mahomed, Birhanu T. Ayele, Charles S. Wiysonge

## Abstract

**Introduction:** The outbreak of novel coronavirus disease 2019 (COVID-19) caught the world off guard in the first quarter of the year 2020. To stem the tide of this pandemic, the development, testing, and pre-licensure approval for emergency use of some COVID 19 vaccine candidates were accelerated. This led to raised public concern about their safety and efficacy, compounding the challenges of vaccine hesitancy which was already declared one of the top ten threats to global health in the year 2019. The onus of managing and administering these vaccines to a skeptical populace when they do become available rests mostly on the shoulders of healthcare workers (HCWs). Therefore, the vaccine confidence levels of HCWs becomes critical to the success of vaccination endeavors, especially COVID 19 vaccination. This proposed study aims to estimate the level of vaccine confidence and the intention to receive a COVID 19 vaccine among future HCWs and their trainers at a specific university in Cape Town, South Africa, and to identify any vaccination concerns early for targeted intervention.

**Methods and analysis:** An online survey will be distributed to current staff and students of an academic institution for HCWs. The survey questionnaire will consist of a demographic questions section consisting of six items and a vaccine confidence section comprising six items in Likert scale format.

A multinomial logistic regression model will be employed to identify factors associated with vaccine confidence and intention. The strength of association will be assessed using odds ratio and its 95% confidence interval. Statistical significance will be defined at a p-value <0.05.

**Ethics and dissemination:** Ethics approval has been obtained for the study from Stellenbosch University (HREC Reference # S19/01/014 (PhD)). The results will be shared with relevant health authorities, presented at conferences, and published in a peer-reviewed journal.

**ARTICLE SUMMARY:** *Strengths and limitations of this study:*

▸ The proposed study will generate baseline knowledge of the vaccine confidence among future healthcare workers and their trainers in its specific context.
▸ It will contribute to addressing the knowledge gap about the intention to receive a COVID 19 vaccine among health care workers in Africa.
▸ It will enable the early identification of vaccine concerns of healthcare workers while they are still in training and assist in informing tailored measures to address them.
▸ A limitation of the study is the possibility of a low response rate which is an inherent challenge of online surveys.

## INTRODUCTION

### Background

The world is currently in the grip of coronavirus disease 2019 (COVID 19) pandemic caused by severe acute respiratory syndrome coronavirus 2 (SARS-CoV-2)[1]. The disease has claimed millions of lives and wreaked economic havoc worldwide since it was declared a pandemic by the World Health Organization (WHO) in March 2020. The problem is further compounded by some countries experiencing a second and third wave of the pandemic[2, 3] and the emergence of new and possibly more virulent strains of the virus[4]. All these, coupled with its high infectivity rate resulted in an unprecedented global effort to develop and produce safe and effective vaccines within an equally unprecedented short time frame[5]. This feat was achieved in the last quarter of the year 2020 when the first set of safe and effective vaccine candidates received pre-licensure emergency use authorization by the Food and Drug Agency in the United States of America and other countries[6].

Since then, there has been a gradual roll out of COVID 19 vaccination with several vaccines by multiple countries. Millions of people have been vaccinated, and many more are scheduled to receive COVID 19 vaccination, raising hopes that the pandemic will soon be brought under control and life can go back to normal.

However, the rapid positive progress in the fight against the COVID 19 pandemic has also brought new concerns. One such concern is the effect of the unprecedented speed of development and approval for use of the successful vaccine candidates on the already tenuous public confidence in vaccines and vaccinations. Prior to the pandemic, the waning public confidence in vaccines that had been simmering for decades reached crisis levels, prompting the WHO to declare ‘vaccine hesitancy’ as one of the ten threats to global health in the year 2019[7]. Defined in 2012 as the delay in acceptance or refusal of vaccination despite the availability of vaccination services[8], vaccine hesitancy is complex and context specific, and varies across time, place and vaccines[8, 9]. Vaccine confidence is one of the most common determinants of vaccine hesitancy globally[10]. Vaccine confidence refers to trust in the safety and effectiveness of vaccines, trust in the competency of the healthcare providers that administer them, and trust in the intentions of the policy makers that propose them[11]. Therefore, estimating the vaccine confidence levels in a population should give an indication of how hesitant or otherwise the population maybe. It has been indicated that an inverse relationship exists between vaccine confidence and vaccine hesitancy[12], therefore an estimate of the level of vaccine confidence would indirectly be indicative of the level of vaccine hesitancy in a given population. This takes on a greater importance if the vaccine confidence investigation is conducted among the subset of the population charged with the responsibility of administering and promoting vaccines and vaccination to the rest of the general public; that is, healthcare workers and their trainers.

The crucial role of healthcare workers to the success of vaccination uptake is well recognized as they are usually the most trusted source of health information for the general public[13–15]. Negative vaccine sentiments expressed by some of them is a cause for concern as those self-reporting to be vaccine hesitant also admit to not administering, recommending, or following the recommended vaccination schedule with their patients[13, 15]. This can undermine vaccination uptake in populations served by such healthcare workers, leading to pockets of under-vaccinated individuals who could potentially serve as reservoirs for outbreaks of vaccine preventable diseases. An undesirable situation to be avoided as much as possible especially in this pandemic period.

### Study rationale

Vaccine attitudes of healthcare workers have been investigated in diverse contexts and places[14, 16–19], and similar studies have been carried out among medical students and healthcare workers in training[20–25]. However, few studies have reported on vaccine attitudes among future health care workers in training and their trainers together. This underscores the need for such a study as this that proposes to estimate the vaccine confidence index among both subsets of the population. In addition, the study will also be providing insight into intention to take COVID 19 vaccines in the study population when they become available. This will be a crucial piece in the knowledge puzzle of the vaccination attitudes among this subset of the population and contribute to that of the wider South African population. In the planned three-phase roll out of COVID 19 vaccination in the country[26], frontline healthcare workers are the priority population scheduled to receive the vaccines in the first phase. Having this baseline knowledge will assist in identifying possible barriers and facilitators to the receipt of the COVID 19 vaccines in the study population, and will provide more recent and supplementary information on intention to take COVID 19 vaccines of the general South African population in addition to those previously reported[10, 27].

In a 67-country survey study of the state of vaccine confidence conducted by Larson and her team 2015, they found that vaccine sentiments were generally positive across the countries surveyed which included South Africa[10]. However, they also reported a diversity of negative sentiments regarding vaccine safety and other issues which varied between continents and countries. The study had four statements related to attitudes towards vaccines: “vaccines are important for children to have”; “overall I think vaccines are safe”; “overall I think vaccines are effective”; and “vaccines are compatible with my religious beliefs”. Participants were asked to rate the degree to which they agree with each statement on the five-point Likert scale: “strongly agree”, “tend to agree”, “do not know”, “tend to disagree,” or “strongly disagree”. The proportion of South African participants who were skeptical of the importance, safety, effectiveness, and religious compatibility of vaccines were 6.6%, 10.2%, 10.2%, and 14.2% respectively. Over half a decade has elapsed since this survey was conducted, the world at large has changed and so has the vaccination landscape, especially with the advent of the severe acute respiratory syndrome corona virus 2 (SARS-CoV-2) and its disease sequalae, the coronavirus disease 2019 (COVID 19) pandemic.

Therefore, it is important and imperative to explore what changes might have occurred over this period of time and what effect it might have on vaccine confidence of South Africans as the country prepares for its COVID-19 vaccination rollout of which the healthcare workers (HCWs) are the scheduled priority recipients in phase 1[26].

HCWs in training are future healthcare practitioners, trainers, and policy makers. As such their vaccine attitudes need to be measured and optimum attention given to them while they are still in training to ensure that a positive vaccination attitude is developed and maintained during their training that will hopefully last long into their professional lives. Therefore, measuring their vaccine confidence is critical to developing targeted interventions that will address potential vaccine concerns that they may be harboring. The attitudes of those who provide training to these future healthcare practitioners are also key to any interventions which seek to foster positive attitudes to vaccines amongst this group. After due consideration of all the cogent reasons proffered above, and the potential benefits inherent in this proposed study, the need for such a study at such a time as this cannot be overemphasized.

### Study aim

The proposed study seeks to answer the following research question: what is the level of vaccine confidence of a cohort of future healthcare workers and their trainers at an academic institution in Cape Town, South Africa? The aim is to estimate the vaccine confidence level of current academic staff and students of the Faculty of Medicine and Health Sciences at a University in Cape Town.

## METHODS AND ANALYSIS

### Study design

This is a cross sectional study including three subsets of the academic institution’s population; academic staff, students, and staff that are also currently pursuing an academic qualification. The operational definition of “academic staff” for the purpose of this proposed study is staff that are engaged in teaching at both undergraduate and post graduate levels. Voluntary participation in the survey will be deemed as consent. Only current academic staff and students are eligible to participate.

### Data collection

The survey will be conducted online using REDCAP survey software to capture participants’ responses. The target population is the entire current academic staff and students of the Faculty of Medicine and Health Sciences of the selected University (i.e. a census of all three sub-groups). It is expected that enough data would be generated through the responses received to power the study to answer the research question. A succinct questionnaire consisting of two types of questions; (1) demographic questions which would be the covariates of the vaccine confidence analysis which includes, but not limited to age and gender, and (2) the four vaccine confidence statements used in the 67 country survey[10] with the addition of two new statements. The first additional statement is about the importance self-vaccination, and the second additional statement is about COVID 19 vaccination intention. All participants will be encouraged to rate all statements as they apply to them. For each vaccine confidence statement, each participant will be asked to rate the degree with which he or she agrees with it; on a five-point Likert scale: strongly agree, tend to agree, do not know, tend to disagree, strongly disagree. The four vaccine confidence statements from the 67 country study[10] are: “vaccines are important for children to have”; “overall I think vaccines are safe”; “overall I think vaccines are effective”; and “vaccines are compatible with my religious beliefs”. In addition, as earlier indicated, we will add one question on vaccine importance (“vaccines are important for me to have”) and one about vaccine intention (“I will take a Covid-19 vaccine when one becomes available”). The responses will be on the same five-point Likert scale as above. The questionnaire will be in English.

### Data analysis plan

After data collection, each questionnaire will be checked for its completeness. Data entry, cleaning and coding will be done using the REDCAP survey software or Microsoft Excel program and exported to Stata software version 16.1 (College Station, TX) for analysis. Categorical variables will be summarized using frequencies and proportions. Continuous covariates will be presented using mean and standard deviation if normally distributed or using median and Inter Quartile Range (IQR) if not normally distributed.

The Chi-square test (for categorical variables) and student’s t test or ANOVA (for continuous variables) will be used to assess the association between vaccine confidence or intention and potential predictors. Multinomial logistic regression model will be employed to identify factors associated with vaccine confidence and intention. The strength of association of will be assessed using Odds Ratio (OR) and its 95% confidence interval. Statistical significance will be defined at a p-value <0.05. The main outcome to be investigated is the variability in the vaccine sentiment and vaccine intention within and across all groups. This will be done by considering the fraction of respondents that will either agree or disagree with the five statements on immunization and the one statement on “intention to vaccinate” previously described[10]. The “strongly agree” and “tend to agree” responses will be combined to make up the positive vaccine sentiments variable, while the “strongly disagree” and “tend to disagree” responses will be combined to make up the negative vaccine sentiment variable. The “don’t know response” and no response will be removed from the data prior analysis.

## LIMITATIONS

A limitation of this proposed study is a possible low response rate which is an inherent limitation of such online surveys. To mitigate against this, the questionnaire is kept short (a total of twelve items in all), drop down options or multiple-choice answers are provided as appropriate to facilitate obtaining timeous, complete and correct responses. At least two reminders, one per week will be sent to follow-up two weeks after the initial email with the survey link is circulated. In addition, cash incentives of a specified amount will be offered to randomly selected participants, one from each group from among the completed, valid received responses. Weighing method would be used to adjust for nonresponse bias should this challenge be encountered[28].

## ORIGINALITY AND ANTICIPATED IMPACT OF THE STUDY

The originality of this study lies in the fact that to the best of our knowledge, no such study has been conducted among the staff and students of the Faculty of Medicine and Health Sciences at this specific University. Anticipated study impact includes but not limited to; the generation of a baseline knowledge of vaccine confidence level of the study population, identifying vaccine confidence issues that are of concern amongst future health care professionals and their trainers, and all the other benefits previously mentioned. This would assist in laying the foundation for the development of tailored interventions to address such concerns by responsible authorities. Moreover, the knowledge generated by this proposed study would add to the growing body of available literature on the potential issues that future health care workers may face and provide evidence to mitigate them while they are still in training.

## ETHICS AND DISSEMINATION

This study obtained ethics approval from Stellenbosch University in South Africa: Human Research Ethics Committee (HREC) reference # S19/01/014 (PhD).

The study results will be presented at conferences and other relevant and appropriate platforms. It will also be published in a peer-reviewed journal.

## DATA STATEMENT

All data relevant to the study will be included in the original research article, or provided as supplementary files.

## Author contributions

EO led the conceptualization, design, and drafted the protocol. BTA collaborated on the data analysis plan and gave feedback on the manuscript; HM and CSW provided supervisory overview and feedback on the overall study, methodology and the manuscript. This team of four authors give their approval to the publishing of this protocol manuscript.

## Data Availability

All data relevant to the study will be included in the original research article or provided as supplementary files.

## Acknowledgements

The statistical input, constructive feedback and promptness of response of BTA is posthumously acknowledged by the surviving three authors.

## Funding

EO gratefully acknowledges the student bursary provided by the South African Medical Research Council (SAMRC) through its’ Internship Scholarship programme; and for funding the open access publication of this protocol through Cochrane South Africa.

## Competing interests

None declared.

## Patient consent

Not required.

## Patient and public involvement

Participation in the survey will be voluntary, the right to participate or decline, and the offer of incentive will be clearly detailed in the survey invite email. Participation in the survey is deemed as consent.

Patients are not involved in the study.

## Ethics approval

The necessary ethics approval has been obtained from the host institution.

## Provenance and peer review

Not commissioned; externally peer reviewed.

## Open access

This is an open access article distributed in accordance with the Creative Commons Attribution Non Commercial (CC BY-NC 4.0) license, which permits others to distribute, remix, adapt, build upon this work non-commercially, and license their derivative works on different terms, provided the original work is properly cited, appropriate credit is given, any changes made indicated, and the use is non-commercial. See: http://creativecommons.org/licenses/by-nc/4.0/.

## Notes

### Competing Interest Statement

The authors have declared no competing interest.

### Funding Statement

The corresponding author (EO) gratefully acknowledges the student bursary provided by the South African Medical Research Council (SAMRC) through its Internship Scholarship program, and for funding the open access publication of this protocol through Cochrane South Africa.

### Author Declarations

This study obtained ethics approval from Stellenbosch University in South Africa: Human Research Ethics Committee (HREC) reference # S19/01/014 (Ph.D.).

## REFERENCES

[1] Listings of WHO’s response to COVID-19 https://www.who.int/news/item/29-06-2020-covidtimeline (accessed 16 January 2021).

[2] Analytical Report of the First, Second and Third Wave Studies https://www.unicef.org/georgia/media/4736/file/COVID-19-Study-Analytical-Report-1-st-2nd-and-3rd-waves-Eng.pdf#page=12&zoom=100,92,96 (2020).

[3] COVID-19 Second wave in South Africa |NICD https://www.nicd.ac.za/covid-19-second-wave-in-south-africa/ (2020, accessed 16 January 2021).

[4] A New Strain of Coronavirus: What You Should Know |Johns Hopkins Medicine https://www.hopkinsmedicine.org/health/conditions-and-diseases/coronavirus/a-new-strain-of-coronavirus-what-you-should-know (accessed 16 January 2021).

[5] Lurie N, Saville M, Hatchett R, et al. Developing Covid-19 Vaccines at Pandemic Speed. N Engl J Med 2020; 382: 1969–1973.

[6] FDA. Emergency Use Authorization |FDA. Food & Drug Administration https://www.fda.gov/emergency-preparedness-and-response/mcm-legal-regulatory-and-policy-framework/emergency-use-authorization (2020, accessed 16 January 2021).

[7] WHO. Ten threats to global health in 2019. World Health Organisation (WHO) 2019; 1–18.

[8] MacDonald NE, Eskola J, Liang X, et al. Vaccine hesitancy: Definition, scope and determinants. Vaccine 2015; 33: 4161–4164.

[9] Betsch C, Böhm R, Chapman GB. Using Behavioral Insights to Increase Vaccination Policy Effectiveness. Policy Insights from Behav Brain Sci 2015; 2: 61–73.

[10] Larson HJ, de Figueiredo A, Xiahong Z, et al. The State of Vaccine Confidence 2016: Global Insights Through a 67-Country Survey. EBioMedicine 2016; 12: 295–301.

[11] Eskola J, Duclos P, Schuster M, et al. How to deal with vaccine hesitancy? Vaccine 2015; 33: 4215–4217.

[12] Orenstein WA, Gellin BG, Beigi RH, et al. Assessing the state of vaccine confidence in the United States: Recommendations from the national vaccine advisory committee. Public Health Rep 2015; 130: 573–595.

[13] Paterson P, Meurice F, Stanberry LR, et al. Vaccine hesitancy and healthcare providers. Vaccine 2016; 34: 6700–6706.

[14] Karafillakis E, Dinca I, Apfel F, et al. Vaccine hesitancy among healthcare workers in Europe: A qualitative study. Vaccine 2016; 34: 5013–5020.

[15] Verger P, Fressard L, Collange F, et al. Vaccine Hesitancy Among General Practitioners and Its Determinants During Controversies: A National Cross-sectional Survey in France. EBioMedicine 2015; 2: 891–897.

[16] Suryadevara M, Handel A, Bonville CA, et al. Pediatric provider vaccine hesitancy: An under-recognized obstacle to immunizing children. Vaccine 2015; 33: 6629–6634.

[17] MacDonald NE, Dubé E. Unpacking Vaccine Hesitancy Among Healthcare Providers. EBioMedicine 2015; 2: 792–793.

[18] Oria PA, Matini W, Nelligan I, et al. Are Kenyan healthcare workers willing to receive the pandemic influenza vaccine? Results from a cross-sectional survey of healthcare workers in Kenya about knowledge, attitudes and practices concerning infection with and vaccination against 2009 pandemic. Vaccine 2011; 29: 3617–3622.

[19] Le Maréchal M, Collange F, Fressard L, et al. Design of a national and regional survey among French general practitioners and method of the first wave of survey dedicated to vaccination. Med Mal Infect 2015; 45: 403–410.

[20] Wibabara Y, Banura C, Kalyango J, et al. Hepatitis B vaccination status and associated factors among undergraduate students of Makerere University College of Health Sciences. PLoS One 2019; 14: 1–9.

[21] Kernéis S, Jacquet C, Bannay A, et al. Vaccine Education of Medical Students: A Nationwide Cross-sectional Survey. Am J Prev Med 2017; 53: e97–e104.

[22] Gallone MS, Gallone MF, Cappelli MG, et al. Medical students’ attitude toward influenza vaccination: Results of a survey in the University of Bari (Italy). Hum Vaccin Immunother 2017; 13: 1937–1941.

[23] Klewer J, Sasnauskaite L, Pavilonis A, et al. Vaccinations in health care students from Germany, Iran, Lithuania and Spain. In: Vaccinations: Types, Potential Complications and Health Effects, pp. 25–43.

[24] Böhme M, Voigt K, Balogh E, et al. Pertussis vaccination status and vaccine acceptance among medical students: multicenter study in Germany and Hungary. BMC Public Health 2019; 19: .PAG-N.PAG.

[25] Loulergue P, Launay O. Vaccinations among medical and nursing students: Coverage and opportunities. Vaccine 2014; 32: 4855–4859.

[26] All You Need To Know About COVID-19 And Vaccines (pdf Guide) - SA Corona Virus Online Portal. 2021; 13.

[27] Lazarus J V., Ratzan SC, Palayew A, et al. A global survey of potential acceptance of a COVID-19 vaccine. Nat Med. Epub ahead of print 2020. DOI: 10.1038/s41591-020-1124-9.

[28] Prince M. Non-Response Bias. In: Core psychiatry https://www.sciencedirect.com/topics/nursing-and-health-professions/nonresponse-bias/pdf (2012).

